# Beyond the Sentence: Clinical and Social Determinants of Forensic Hospitalization Duration in Northern Israel

**DOI:** 10.64898/2026.06.25.26356525

**Authors:** Ilana Kovalenko, Shiran Simonov, Alon Shamir, Laura Sharony

**Affiliations:** The Azrieli Faculty of Medicine, Bar-Ilan University, Safed, Israel; Psychobiology Research Lab, Mazor Mental Health Center, Akko, Israel; The Ruth and Bruce Rappaport Faculty of Medicine, Technion Israel Institute of Technology, Haifa, Israel

**Keywords:** Length of stay, forensic psychiatry, hospitalization, psychiatric disorder

## Abstract

**Purpose:** Involuntary psychiatric hospitalization under court orders requires careful balancing of legal obligations and clinical needs. Identifying factors that influence the length of these hospital stays helps clarify the relationship between legal frameworks and psychiatric treatment. This study aims to describe the socio-demographic, clinical, and legal profiles of individuals hospitalized under court warrants and to identify factors independently associated with the duration of forensic hospitalization.

**Methods:** A retrospective study was conducted on 119 patients discharged between 2018 and 2023. Data were collected from medical and legal records, including socio-demographic details, psychiatric diagnoses, offense types, hospital stay lengths, and legal proceedings.

**Results:** Most patients were men (91.6%) diagnosed with schizophrenia or schizoaffective disorder (97.5%), with high rates of comorbid substance use disorder (79.0%) and unemployment (85.7%). The median hospital stay was 19.0 months, representing 40% of the maximum statutory sentence. Patients with low-severity offenses served a larger share of their maximum sentence (47%) than those with high-severity offenses (24%). Time to first discretionary leave was the strongest predictor of total stay duration in univariable analysis.

**Conclusion:** The finding that patients with minor offenses have longer hospital stays than those with serious offenses confirms that clinical factors, rather than offense severity, primarily influence discharge decisions. These findings support moving toward personalized, clinically focused, and family-inclusive forensic discharge planning while maintaining public safety.

## Introduction

Psychiatric hospitalization under the Treatment of the Mentally Ill Law (1991) comprises two main categories: voluntary and involuntary. Involuntary hospitalization can be initiated either by the district psychiatrist or through a court-issued order. Courts evaluate a defendant’s mental competence via psychiatric assessments, which may include temporary hospitalization under a “court order for hospitalization supervision.” If the individual is deemed unfit to stand trial or not criminally responsible, the court may issue an “involuntary hospitalization order” or an “involuntary outpatient treatment order” (see Fig. 1). Every six months or sooner, a psychiatric committee reviews the patient’s mental status and determines whether they should be discharged or continue their hospitalization. It is important to note that patients are entitled to legal representation during appeal hearings. [1]

**Figure 1.**
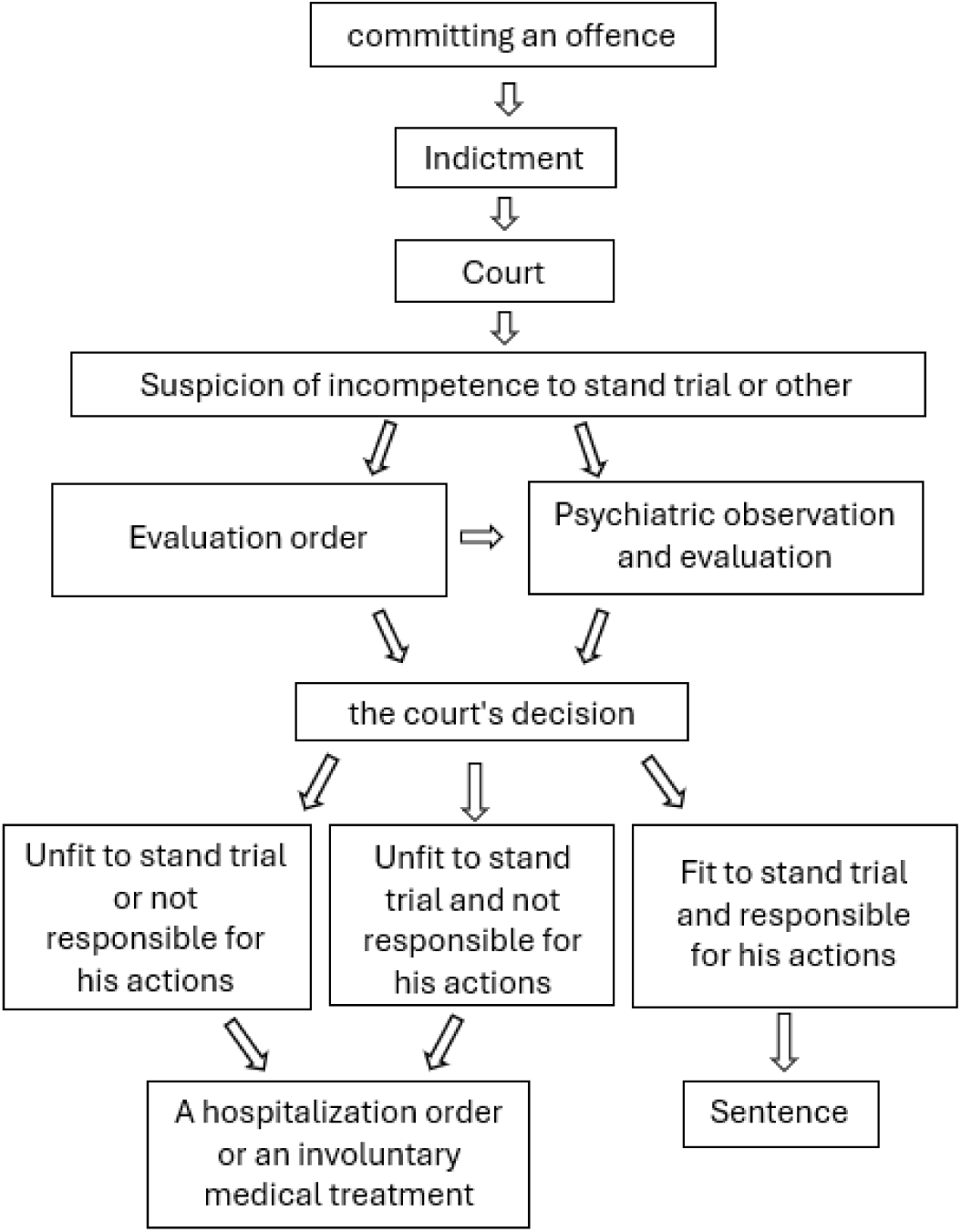
Forensic Psychiatric Evaluation Process in Israel.

Israel has seen significant fluctuations in involuntary psychiatric hospitalizations over the past twenty years. The 2004 Mental Health Report showed a concerning rising trend from 1996 to 2003, with court-ordered involuntary admissions steadily increasing to make up 21.2% of all psychiatric hospitalizations by 2003 [2]. However, recent years have seen a clear reversal of this trend. After peaking at 37% in 2020, involuntary hospitalizations dropped to 34% in 2021 and further fell to 29% of all psychiatric admissions in 2022. This positive change is also evident in population-based rates, with court-ordered involuntary hospitalizations occurring at 0.13 per 1,000 residents in 2022—a 30% decrease from 2021 and a 41% decrease from 2019. Despite these advances, involuntary admissions still account for a significant portion across various admission types, with 30% of first-time admissions and 28% of readmissions being involuntary in 2022. Forensic observation cases showed similar improvements, dropping from 67% of admissions being involuntary in 2019 to 60% in 2021 and 45% in 2022. Diagnostically, schizophrenia and delusional disorders remained the most common conditions among involuntarily hospitalized patients, representing 48% and 43% of cases, respectively. While the recent decline indicates meaningful progress, the overall share of involuntary admissions remains relatively high [3].

Involuntary hospitalization requires balancing patients’ rights to treatment, dignity, and freedom with the need to protect public safety. This balance often creates tension between medical and legal systems, influencing patient outcomes and societal risk. The release process is similarly complex, as court-mandated hospitalization durations are subject to appeal. As a result, patients may face short hospitalizations after severe offenses or extended stays following minor ones, depending on their mental state and legal considerations [4].

Common challenges in involuntary hospitalization include inadequate justification for admission, limited consideration of less restrictive options, and concerns about procedural fairness during psychiatric committee reviews. Additionally, the option to convert hospitalization warrants into outpatient treatment remains underused, despite its potential to shorten hospital stays and reduce rehospitalization rates [2].

Interestingly, 53% of early-career psychiatrists support reevaluating national laws on involuntary hospitalization. Internationally, psychiatrists in high-income countries report more ethical and bureaucratic challenges, while those in low- and middle-income countries tend to report fewer difficulties [1].

A recent survey found that short-term hospitalization increased patient trust in the legal process and improved satisfaction with medication treatment. Importantly, no benefit of long-term hospitalization was seen in recidivism or readmission rates. These results suggest that more extended hospital stays may not meet their therapeutic goals and can lead to practical issues, such as high costs and overcrowding [6].

In Hong Kong, a study of 4,492 criminal defendants referred for psychiatric evaluation between 2011 and 2016 found that most were males aged 18–39, commonly charged with theft, violence, or drug-related offenses, and 73% of cases were diagnosed with schizophrenia, substance use disorders, and affective disorders being the most prevalent [7]. Another study gathered comprehensive socio-demographic and clinical data, including education, employment, ethnicity, income, family and psychiatric history, criminal records, and substance use. Results indicated that 60% of offenders had prior psychiatric hospitalizations, with schizophrenia identified in 66.7% of cases. Additionally, 86.7% had multiple police records, and 93.3% were treated with antipsychotic medication. Nearly half reported adverse childhood experiences, including abuse, substance exposure, parental absence, maternal abuse, or living with a mentally ill family member [8]. Criminal offense, community residential setting into which the patient was released, age, and gender were reported to be significantly associated with inpatient LoS in Hungary [9]. More recently, a cross-sectional survey of 291 forensic inpatients in Hunan, China, confirmed comparable socio-demographic and clinical profiles across cultures, with schizophrenia spectrum disorders and substance use disorder dominating the diagnostic picture [10].

Between 2018 and 2020, the length of forensic stays at Mazor Mental Health Center ranged from 489 to 694 days. Therefore, we conducted a retrospective analysis to characterize the socio-demographic, clinical, and legal profiles of individuals hospitalized under court warrants and identify factors associated with the duration of forensic stays. Here, we express the length of stay as a proportion of the maximum statutory sentence (stay-to-sentence ratio), which provides an analytic framework that applies across legal frameworks by normalizing absolute durations relative to each system’s legal ceiling and thereby exposing clinical rather than legal drivers of discharge.

## Methods

### Subjects

This work was approved by the Helsinki Committee of the Mazor Mental Health Center (01-23-MZR). 119 medical files of discharged forensic inpatients were sampled between 2018 and 2023. The current work did not include medical records of individuals with severe criminal offenses and court orders that were over 20 years.

### Data Collection

Demographic, clinical, socioeconomic, and forensic data were collected (see variables in Supplemental Table 1) and included the following Socio-Demographic characteristics: age, sex, country of birth, living status, marital status, number of children, education, employment, and religion.

Clinical data: diagnosis (schizophrenia / schizoaffective disorder/mood disorder/none of these), personality disorder, organic disorder, substance use disorder. Number of involuntary hospitalizations before the current hospitalization.

Forensic data includes the type of offense, the maximum statutory sentence associated with the offense category, duration of stay, number of committees conducted until the first discretionary leave, length of stay until the first leave, and number of committees until discharge or transfer to compulsory outpatient treatment.

The offenses were categorized into three types: offenses against a person, offenses against property or drugs, and combined offenses. Offenses against a person included violent crimes (such as offenses against human life, the human body, assault of any kind, or arson), offenses with the potential for violence (such as violation of protection and restriction orders or threats), and sexual offenses. Offenses against property encompass property and drug crimes (damage to property, break-ins, thefts, drug possession and trafficking), economic crimes (fraud, financial offenses, licensing violations, and bribery), and traffic violations.

### Statistical Analysis

Given the non-normal distribution of all continuous variables, confirmed by Shapiro–Wilk tests (all p<.001, Supplementary Table 1), non-parametric statistical methods were applied throughout. Participants were stratified into two complementary comparison frameworks: (i) by actual hospitalization length, using tertile-based cutoffs to create short- (≤14 months), medium- (15–24 months), and long-stay (≥25 months) groups; and (ii) by offense severity, operationalized as the maximum statutory sentence length divided into tertile-based groups (low: ≤36 months; medium: 37–60 months; high: >60 months).

Between-group differences in continuous variables were assessed using the Kruskal–Wallis H test, with Dunn–Bonferroni post hoc comparisons for pairwise testing. Effect sizes were quantified using epsilon-squared (ε²), where 0.01, 0.06, and 0.14 correspond to small, medium, and large effects, respectively. Chi-square tests with Cramér’s V were used for categorical variables. Fisher’s exact test was applied when expected cell counts were below 5. Monotonic associations between continuous variables were quantified using Spearman’s rank correlation (ρ). Mann–Whitney U tests were used for two-group comparisons.

To identify independent predictors of hospitalization duration, multiple linear regression was performed with log-transformed stay duration as the dependent variable to address positive skewness (R² = .333, Adjusted R² = .271, F = 5.39, p < .001). Separately, logistic regression with L2 penalization identified predictors of prolonged stay, defined as ≥25 months (the upper tertile), with confidence intervals derived from 500-sample bootstrap resampling to address quasi-separation due to the limited sample size. All statistical analyses were performed using Python (version 3.12).

## Results

This study reviewed 119 medical records of patients hospitalized under court-ordered warrants and discharged between 2018 and 2023. Cases with incomplete primary outcome data or non-qualifying diagnoses were excluded from specific sub-analyses as described below.

### Socio-demographic Data

The socio-demographic profile of patients hospitalized under court orders at Mazor Mental Health Center showed that most patients were single (67.2%), unemployed (85.7%), male (91.6%), with a mean age of 42.3 years (SD = 12.4). The majority were born in Israel (74.8%), followed by individuals from the former Soviet Union (17.6%) and other countries (7.6%). Over half (57.1%) lived with their families, and the majority had a high school education or below (67.9%). The primary religious affiliation was Jewish (55.5%), with minorities including Muslims (29.4%), Druze (8.4%), and Christians (3.4%) (Table 1).

**Table 1.** Social and demographic characteristics of patients hospitalized under court order at Mazor Mental Health Medical Center.

| Characteristics |  | N | (%) |
| --- | --- | --- | --- |
| Sex | Male | 109 | 91.6 |
|  | Female | 10 | 8.4 |
| Marital status | Single | 80 | 67.2 |
|  | Divorced | 20 | 16.8 |
|  | Widowed | 1 | 0.8 |
|  | Married | 18 | 15.1 |
| Education | No matriculation certificate | 8 | 6.7 |
|  | High school | 73 | 61.3 |
|  | University degree | 33 | 27.7 |
|  | Unknown | 5 | 4.2 |
| Living status | Family | 68 | 57.1 |
|  | Alone | 51 | 42.9 |
| Country of birth | Israel | 89 | 74.8 |
|  | Former Soviet Union | 21 | 17.6 |
|  | Other | 9 | 7.6 |
| Employment | Employed | 17 | 14.3 |
|  | Unemployed | 102 | 85.7 |
| Religion | Jewish | 66 | 55.5 |
|  | Muslim | 35 | 29.4 |
|  | Druze | 10 | 8.4 |
|  | Christian | 4 | 3.4 |
|  | Unknown | 4 | 3.4 |

### Clinical Data

The primary admission diagnosis was an acute psychotic condition. As shown in Table 2, 71.4% of patients were diagnosed with schizophrenia, 26.1% with schizoaffective disorder, and 0.8% with a mood disorder. Comorbid personality disorders were present in 42.0% of cases, and organic disorders in an additional proportion of patients. Substance use disorder was documented in 79.0% of patients. Most patients (63.9%) had no prior involuntary psychiatric hospitalization at the time of the index admission; 20.2% had one prior admission, 10.9% had two, and 5.0% had three (Table 2).

**Table 2.** Clinical and Psychiatric Characteristics of Court-Ordered Forensic Inpatients.

| Psychiatric Characteristics |  | N | (%) |
| --- | --- | --- | --- |
| Psychiatric diagnosis | schizophrenia | 85 | 71.4 |
|  | schizoaffective | 31 | 26.1 |
|  | mood disorder | 1 | 0.8 |
|  | None of the above | 2 | 1.7 |
| Personality disorder | yes | 50 | 42 |
|  | no | 69 | 58 |
| Drug use | yes | 94 | 79 |
|  | no | 25 | 21 |
| Number of involuntary hospitalizations before the current hospitalization | 0 | 74 | 62.2 |
|  | 1 | 24 | 20.7 |
|  | 2 | 12 | 10.3 |
|  | 3 | 6 | 5.2 |

### Offense Types

Patient offenses were categorized as against a person (63.9%), property or drug-related (5.0%), or combined offenses (31.1%). Maximum statutory sentences ranged from 18 to 240 months, with a median of 36 months (mean 56.6, SD= 38.6). A Kruskal–Wallis test revealed no statistically significant difference in maximum sentence length across offense type categories (H= 2.17, p=.34) (Supplemental Table 1).

### Length of Stay

Lengths of stay ranged from 6 to 97 months (median= 19.0 months, IQR 13.5–32.2; mean= 23.1, SD= 14.5). Most patients (84.9%) served only part of their mandated hospitalization period. Relative to the maximum statutory sentence for their offense category, actual stays represented a median of 40% (mean 48.3%, SD= 28.4) of the maximum sentence.

### Comparison by Hospitalization Length Group

Patients were stratified into three groups by length of hospitalization: short stay (≤14 months; n= 40), medium stay (15–24 months; n= 42), and long stay (≥25 months; n= 37). Group comparisons revealed a significant gradient in the maximum statutory sentence (short: median 36 months, medium: 48 months, long: 60 months; H= 12.82, p=.002, ε²=.093). The ratio of actual stay to maximum sentence increased markedly across groups (short: 30%, medium: 40%, long: 70%; H= 32.83, p<.001, ε²=.266), indicating that long-stay patients served a substantially larger proportion of their maximum statutory sentence. The proportion of patients completing the full court-ordered hospitalization period also increased (0% short, 14% medium, 32% long; χ²= 15.78, p<.001, V=.364).

Time to first discretionary leave showed the largest effect size among all analyses, increasing sharply across groups: median 7.0 months [IQR 6–8] for the short-stay group, 10.8 months [7–15] for the medium-stay group, and 23.0 months [18–29] for the long-stay group (H = 47.52, p<.001, ε²= .474) (Table 3). The rate of transfer to compulsory outpatient treatment decreased significantly across groups (short: 100%, medium: 86%, long: 65%; χ²= 17.82, p<.001, V= .387). Family attitudes toward discharge also differed significantly. Ambivalent or opposing family positions were much more common in the long-stay group (41%) than in the short-stay group (5%; χ²= 20.14, p<.001, V=.291). Lawyer absence at committee hearings was significantly more frequent among long-stay patients (16%) compared to short-stay patients (0%; χ²= 10.57, p=.005, V=.298). Female sex was also significantly linked to shorter stays (short group: 18% female vs. long group: 3%; χ²= 6.59, p=.037, V= .235).

**Table 3.**
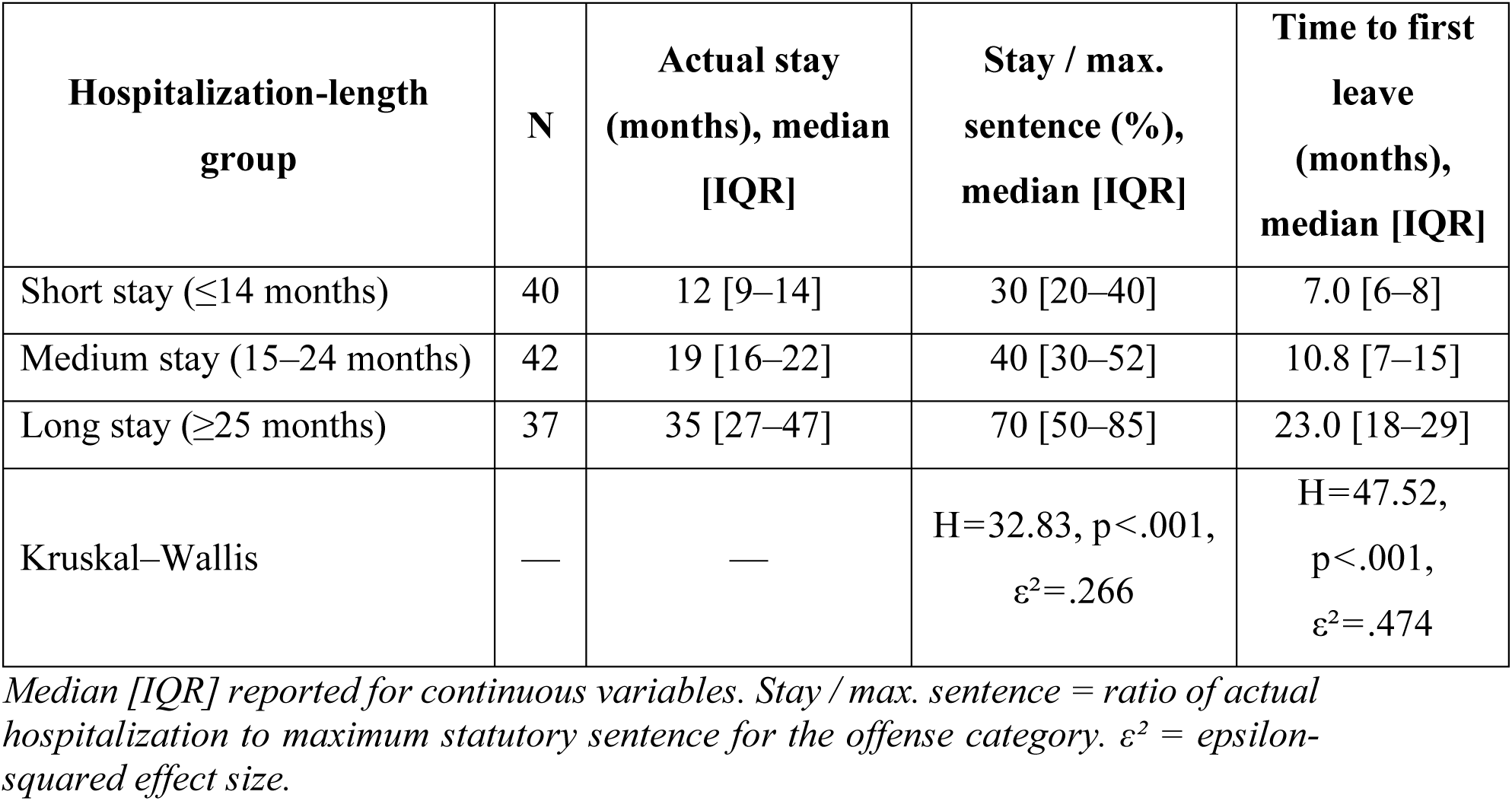
Length of stay and key process indicators by hospitalization-length tertile group.

| Hospitalization-length group | N | Actual stay (months), median [IQR] | Stay / max. sentence (%), median [IQR] | Time to first leave (months), median [IQR] |
| --- | --- | --- | --- | --- |
| Short stay ($\leq 14$ months) | 40 | 12 [9–14] | 30 [20–40] | 7.0 [6–8] |
| Medium stay (15–24 months) | 42 | 19 [16–22] | 40 [30–52] | 10.8 [7–15] |
| Long stay ($\geq 25$ months) | 37 | 35 [27–47] | 70 [50–85] | 23.0 [18–29] |
| Kruskal–Wallis | — | — | H=32.83, $p < .001$, $\epsilon^2 = .266$ | H=47.52, $p < .001$, $\epsilon^2 = .474$ |

*Median [IQR] reported for continuous variables. Stay / max. sentence = ratio of actual hospitalization to maximum statutory sentence for the offense category. $\epsilon^2$ = epsilon-squared effect size.*

### Comparison by Offense Severity

When patients were stratified by offense severity using maximum statutory sentence (low: ≤36 months, n= 66; medium: 37–60 months, n= 24; high: >60 months, n= 29), actual hospitalization duration increased monotonically with severity (low: median 14.8 months [IQR 12–24], medium: 19.5 [16–29], high: 24.0 [16–35]; H= 10.98, p=.004, ε²= .077). In contrast, the stay-to-maximum-sentence ratio showed the inverse pattern, decreasing with increasing severity (low: 47%, medium: 32%, high: 24%; H= 24.88, p<.001, ε²= .197). This inverse relationship confirms that patients with the most serious offenses remain hospitalized longer in absolute terms but serve a proportionally smaller share of their maximum statutory sentence. Time to first discretionary leave also lengthened significantly with increasing severity (low: 8.0 months [6–10], medium: 13.0 [10–19], high: 15.0 [10–25]; H= 16.96, p<.001, ε²= .156), and high-severity patients were less likely to be transferred to compulsory outpatient treatment (low: 74%, medium: 96%, high: 97%; χ²= 10.59, p=.005, V= .298) (Table 4).

**Table 4.** Hospitalization process indicators by offense severity group (maximum statutory sentence tertiles)

| **Variable** | **Low severity**<br>**(≤36 months)**<br>**n=66** | **Medium**<br>**severity (37–**<br>**60 months)**<br>**n=24** | **High severity**<br>**(>60 months)**<br>**n=29** | **p-value** |
| --- | --- | --- | --- | --- |
| Actual stay (months), median [IQR] | 14.8 [12–24] | 19.5 [16–29] | 24.0 [16–35] | .004 |
| Stay / max. sentence, median (%) | 47 | 32 | 24 | <.001 |
| Time to first leave (months), median [IQR] | 8.0 [6–10] | 13.0 [10–19] | 15.0 [10–25] | <.001 |
| Total committees until discharge, median [IQR] | 2.0 [1–2] | 3.0 [2–3] | 2.0 [2–3] | <.001 |
| Transferred to outpatient treatment, n (%) | 49 (74%) | 23 (96%) | 28 (97%) | .005 |
| Full hospitalization (not conditional), n (%) | 17 (26%) | 0 (0%) | 1 (3%) | .001 |

*p-values from Kruskal–Wallis $H$ test (continuous) or chi-square test (categorical). Median [IQR] for continuous variables. Low: ≤36 months; Medium: 37–60 months; High: >60 months.*

### Effect of Socio-demographic Parameters on Hospitalization Length

No significant correlation was found between patient age and length of hospitalization (Spearman ρ = .029, p = .753). A significant negative correlation was observed for the number of children (ρ = –.199, p= .030), indicating that patients with more children tended to have shorter stays. Female sex was significantly and negatively correlated with stay duration (ρ =–.260, p= .004). Other sociodemographic variables, including living situation, employment, marital status, education level, religion, and country of birth, showed no significant correlation with hospitalization duration (Supplemental Table 2).

### Committees During Hospitalization

During hospitalization, most patients underwent one or more committee evaluations for discretionary leave, a hospital-approved temporary absence typically granted to patients demonstrating clinical improvement. Of 119 patients, 20 (16.8%) did not receive discretionary leave, while 99 (83.2%) received approval for leave. The median waiting period for leave approval was 10.0 months [IQR 6–17] (mean= 12.7, SD= 8.1, range 4–46 months). Overall, 100 patients (84.0%) were transferred to compulsory outpatient treatment.

Spearman’s correlations revealed no significant association between total committee count and hospitalization duration (ρ = – .140, p = .128). However, among patients transferred to compulsory outpatient treatment (n= 100), the number of release committees showed a significant negative correlation with length of stay (ρ =–.270, p= .003), suggesting that more frequent review hearings facilitate earlier discharge. The strongest single predictor was time to first discretionary leave (ρ =.743, p<.001, n= 99), confirming that early leave approval strongly predicts the total treatment course (Supplemental Table 2).

Time to first leave differed significantly across both offense severity groups (H = 16.96, p<.001, ε²=.156) and hospitalization-length groups (H = 47.52, p<.001, ε²=.474), making it the single most discriminating indicator across both comparison frameworks.

### Legal Course by Psychiatric Diagnosis

Patients were categorized into diagnostic groups: schizophrenia (n= 85, 71.4%), schizoaffective disorder (n= 31, 26.1%), mood disorder (n= 1, 0.8%), and other (n= 2, 1.7%). Mood disorders and unclassified cases were excluded from comparisons of diagnostic groups. No statistically significant differences were found in offense severity distribution across diagnostic groups (χ²(6) = 6.62, p=.36).

Patients with schizophrenia had significantly longer hospitalizations before transfer or discharge (median 20.0 months [IQR 14–34]) compared to patients with schizoaffective disorder (median 14.5 months [IQR 12–22]; Mann–Whitney U= 1727, p=.011). Time to first leave approval did not differ significantly between these diagnostic groups (schizophrenia: median 10.0 months, schizoaffective: 8.0 months; p= .115), nor did offense severity distribution (χ²(2) = 4.87, p= .088).

### Predictors of Hospitalization Duration

Univariable Spearman correlations identified the following significant predictors of longer stay: time to first leave (ρ =+.743, p< .001), transfer to outpatient treatment (ρ =– .445, p< .001), maximum statutory sentence (ρ =+.293, p=.001), lawyer presence at committee (ρ =–.291, p= .001), number of release committees (ρ = –.270, p=.003), receipt of leave (ρ =–.261, p= .004), female sex (ρ =–.260, p=.004), number of children (ρ =–.199, p=.030), and family opposition to discharge (ρ =+.188, p= .040).

In the multiple linear regression model with log-transformed stay duration as the dependent variable (N= 119, R²= 0.333, Adjusted R²= 0.271, F= 5.39, p< .001), three predictors were independently significant (Table 5). Maximum statutory sentence length was the strongest predictor (β=+.006, SE=.001, t= 4.64, p<.001). Each additional month of statutory sentence was independently associated with longer actual stay duration. Female sex was independently associated with markedly shorter hospitalization (β=–.478, SE=.160, t=–.299, p=.003), even after adjusting for offense severity and all other covariates. Family opposition to discharge was also a significant independent predictor of longer stay (β=+.515, SE=.244, t= 2.11, p=.037). Lawyer presence at committee hearings showed a marginal trend toward association with shorter stay (β=–.397, SE=.221, t=–.180, p= .074). Substance use disorder, personality disorder, prior admissions, age, total committees, and court appeal status did not independently predict hospitalization length after covariate adjustment (all p> .10).

**Table 5.** Multiple linear regression: predictors of log-transformed hospitalization duration (N = 119, R²= 0.333, Adj-R²= 0.271)

| Predictor | $\beta$ | SE | t | P-value | 95% CI |
| --- | --- | --- | --- | --- | --- |
| Intercept | 3.132 | 0.265 | 11.81 | <.001 | [2.612, 3.652] |
| Maximum statutory sentence (months) | 0.006 | 0.001 | 4.64 | <.001 | [0.003, 0.008] |
| Female sex | -.478 | 0.160 | -2.99 | .003 | [-0.791, -0.165] |
| Family against discharge | 0.515 | 0.244 | 2.11 | .037 | [0.036, 0.994] |
| Lawyer present at committee | -.397 | 0.221 | -1.80 | .074† | [-0.830, 0.035] |
| Prior forensic admissions | 0.007 | 0.053 | 0.13 | .899 | [−0.098, 0.111] |
| Age (years) | 0.001 | 0.004 | 0.35 | .729 | [−0.006, 0.009] |
| Total committees | —<br>0.087 | 0.057 | −1.52 | .132 | [−0.200, 0.025] |
| Substance use disorder | 0.084 | 0.116 | 0.73 | .470 | [−0.143, 0.312] |
| Personality disorder | —<br>0.008 | 0.096 | −0.08 | .933 | [−0.196, 0.180] |
| Filed a court appeal | —<br>0.230 | 0.296 | −0.77 | .440 | [−0.810, 0.351] |

$\beta$ = unstandardized coefficient for $\log(\text{stay duration in months})$ . A positive $\beta$ indicates association with longer stay. † $p < .10$ ; \* $p < .05$ ; \*\* $p < .01$ .

## Discussion

This retrospective study examined the socio-demographic, clinical, and legal profiles of 119 forensic psychiatric inpatients hospitalized under court orders at Mazor Mental Health Center and identified factors independently associated with the duration of hospitalization. Using non-parametric methods appropriate for the right-skewed distribution of stay duration, and employing two complementary grouping frameworks by hospitalization length and by offense severity, we identified several clinically and legally meaningful predictors.

The principal finding was the inverse relationship between offense severity and the proportion of court-mandated time served. Patients with low-severity offenses served a median of 47% of their maximum statutory sentence, compared to 32% for medium-severity and only 24% for high-severity offenders (H = 24.88, p<.001, ε²=.197). These data confirm that psychiatric committees prioritize clinical readiness for discharge over adherence to the legal ceiling imposed by the sentence, consistent with the framework of the Israeli Mental Health Act and prior findings from Israeli forensic facilities (1,2,4). Regardless of sentence length, patients require a minimum period of hospitalization to stabilize psychiatrically, reduce risk, and build functional recovery capacity. The finding that patients with minor offenses serve a disproportionate amount of their court-ordered time reflects that the minimum clinically required period constitutes a larger fraction of their shorter maximum sentence.

Time to first discretionary leave was the strongest predictor of total hospitalization duration. This finding is consistent with clinical experience. Early leave approval signals early recovery and initiates the trajectory toward discharge. It supports the use of systematic early clinical review and structured leave protocols within the first six to twelve months of forensic admission. The high rates of leave approval (83.2%) and transition to compulsory outpatient treatment (84.0%) confirm the broad feasibility of community-based forensic care and align with the underutilized recommendation to convert to outpatient treatment [2]. Recent evidence confirms that supervised compulsory outpatient treatment consistently reduces criminal recidivism and improves outcomes for forensic patients worldwide, while unconditional discharge without community supervision is linked to significantly higher reoffending rates [11].

The multivariate regression model identified three significant independent predictors of length of hospitalization. The maximum statutory sentence was the strongest, reflecting the expected legal influence on stay durations. Family opposition to discharge was the second significant predictor. Although only 3.4% of families were explicitly recorded as opposing discharge, a further 26.1% held ambivalent, and this combined non-supportive posture was strongly over-represented in the long-stay group (41%) relative to the short-stay group. This finding underscores the underappreciated role of family dynamics in forensic discharge planning and identifies systematic family engagement and psychoeducation as potentially modifiable targets for reducing stay duration [5]. Research demonstrates that family involvement during psychiatric inpatient care is associated with more comprehensive discharge planning and earlier initiation of outpatient follow-up [12]. In forensic settings specifically, barriers to family-centered care remain entrenched [13], and a recent systematic review found that families of inpatients consistently report exclusion from treatment decisions, with collaborative discharge planning dissatisfied by institutional constraints on communication [14].

Notably, female sex was a significant independent predictor of shorter stay, suggesting that women face meaningfully different clinical trajectories or risk-weighting by committees, even after adjusting for offense severity.

Additionally, the presence of a lawyer at committee hearings showed a marginal independent association with shorter stay. Although it does not meet conventional significance thresholds, this finding is consistent with the Spearman correlation and warrants further investigation. Legal representation may facilitate more structured committee proceedings, improve advocacy for clinically appropriate discharge, or signal broader social support. The low overall rate of missing legal representation (only 16% in the long-stay group and 0% in the short-stay group) also highlights a systemic inequity that deserves clinical attention.

Substance use disorder, present in 79.0% of patients, at the higher end of the 50–80% prevalence range reported internationally [15,16], did not independently predict longer stay in the present analysis (p= .470). This may reflect the high and relatively uniform prevalence of SUD across the cohort, limiting its discriminatory power within this sample. Nevertheless, its high prevalence is clinically significant given the established relationship between SUD, violence, and mental illness severity in forensic settings [19,20].

Similar to international studies [7,8], the cohort was predominantly male, unemployed, single, and diagnosed with schizophrenia spectrum disorders. Most patients (63.9%) had no prior forensic hospitalization, suggesting potential gaps in early community treatment or identification before the index offense. Patients with schizophrenia had significantly longer hospitalizations than those with schizoaffective disorder, likely reflecting the greater burden of negative symptoms and slower functional rehabilitation associated with schizophrenia [21,22]. This difference should be interpreted with caution, given the smaller schizoaffective group.

Beyond Israel-specific implications, these findings contribute to the cross-national epidemiology of service systems in forensic psychiatric care. Reported lengths of stay in forensic and involuntary psychiatric care differ markedly across jurisdictions, ranging from a mean of 23.8 days in a Greek general-psychiatric involuntary cohort to medians of 18–39 months in Polish, Hungarian, and Swiss forensic samples (9, 23). Absolute durations are therefore difficult to interpret without reference to the underlying legal ceiling. Expressing duration as a proportion of the maximum statutory sentence, as done here, partially controls for these jurisdiction-specific differences and exposes the clinical drivers of discharge that absolute durations obscure. The inverse pattern we report, proportionally longer stays for minor offenses, may therefore represent a system-level signature of clinically (rather than legally) governed discharge and warrants replication in other forensic services that operate under similar dual mandates of treatment and public protection.

### Strengths and Limitations

Mazor Mental Health Center serves a genuinely multi-ethnic population, including Jewish, Muslim, Druze, and Christian patients. This diversity enhances the ecological validity of the socio-demographic findings and broadens their applicability beyond mainly monoethnic forensic settings. The study examined variables across four distinct areas (socio-demographic, clinical, legal, and committee process), enabling a more thorough characterization of predictors than studies that rely solely on administrative or legal data. Including committee process variables such as time to first discretionary leave, committee count, alignment, and family status captures aspects of the hospitalization trajectory that diagnosis and offense type alone cannot. Using two complementary analytical frameworks, grouping patients by hospitalization length and by offense severity, provides triangulated evidence supporting the key finding that clinical readiness, rather than legal sentence length, determines discharge decisions. Methodologically, the introduction of the stay-to-sentence ratio offers a transferable normalization device for forensic length-of-stay research that is not confounded by jurisdiction-specific differences in the legal ceilings of offense categories, enabling more meaningful international comparison than absolute durations allow.

Several limitations must be acknowledged. The retrospective single-center design limits causal inference and generalizability. Cross-national comparisons are complicated by variation in legal frameworks, security levels, and service configurations, as demonstrated by recent studies from Switzerland [23], Germany [24], and Hungary (See introduction). The relatively small sample (N = 119) reduces statistical power for subgroup analyses and for reliable estimation of rare predictors, including court appeals (n= 3, 2.5%) and explicit family opposition (n= 4, 3.4%). This finding should therefore be interpreted as directionally consistent and clinically plausible rather than definitively established. The exclusion of court orders more than 20 years old may attenuate associations at the extreme end of offense severity. The predominantly male composition restricts the interpretation of sex-related findings. Future studies should incorporate standardized actuarial risk instruments, family assessment measures, and multi-center designs to validate and extend these findings.

Although the observation-to-predictor ratio in our principal regression is within accepted ranges, secondary coefficient estimates, particularly those involving low-prevalence categorical predictors, should be interpreted as directional, with the reported bootstrap-derived confidence intervals (rather than the point estimates alone) treated as the relevant measure of precision.

## Conclusion

This study demonstrates the complex interplay of legal, clinical, and social factors that determine the duration of forensic psychiatric hospitalization. The inverse relationship between offense severity and proportional stay time confirms that clinical readiness for discharge, not the legal ceiling of the sentence, primarily drives committee decisions. Time to first leave approval is the most powerful univariable predictor of total stay, underscoring the value of structured early clinical assessment. The high rate of successful transition to outpatient treatment (84.0%) confirms the feasibility of community-based forensic care for most patients. These results support a shift toward personalized and clinically focused care. Further prospective research incorporating standardized clinical risk instruments is needed to clarify how committee decision-making integrates these factors.

## Data Availability

All data produced in the present study are available upon reasonable request to the authors

## Funding and/or Competing interests

The authors have no relevant financial or non-financial interests to disclose.

## Funding

This research did not receive any specific funding from agencies in the public, commercial, or not-for-profit sectors.

## Notes

### Competing Interest Statement

The authors have declared no competing interest.

### Author Declarations

This work was approved by the Helsinki Committee of the Mazor Mental Health Center (01-23-MZR).

